# Safety, Immunogenicity, and Efficacy of COVID-19 Vaccine in Children and Adolescents: A Systematic Review

**DOI:** 10.1101/2021.09.11.21262855

**Authors:** Meng Lv, Xufei Luo, Quan Shen, Ruobing Lei, Xiao Liu, Enmei Liu, Qiu Li, Yaolong Chen

## Abstract

**Aim:** To identify the safety, immunogenicity, and protective efficacy of COVID-19 vaccine in children and adolescents.

**Methods:** We conducted a systematic review. Databases including PubMed, Web of Science, WHO COVID-19 database, and CNKI were searched on 23 July 2021. International Clinical Trials Registry Platform (ICTRP) was also searched to collect ongoing trials. We included published researches or ongoing clinical trials related to the safety, immunogenicity, and efficacy of COVID-19 vaccine in children or adolescents (aged ≤18 years). Meta-analysis was performed if the consistency of the included studies was high. If not, descriptive analyses were performed.

**Results:** Eight published studies with 2851 children or adolescents and 28 ongoing clinical trials were included. Among eight published studies, two (25.0%) were RCTs, two (25.0%) case series, and four (50.0%) case reports. The results showed selected COVID-19 vaccines had a good safety profile in children and adolescents. Injection site pain, fatigue, headache, and chest pain were the most common adverse events. Some studies reported a few cases of myocarditis and pericarditis. Two RCTs showed that the immune response to BNT162b2 in adolescents aged 12-15 years was non-inferior to that in young people aged 16-25 years, while a stronger immune response was detected with 3μg CoronaVac injection. Only one single RCT showed the efficacy of BNT162b2 was 100% (95% CI: 75.3 to 100). Of the 28 ongoing clinical trials, twenty-three are interventional studies. Fifteen countries are conducting interventional clinical trials of COVID-19 vaccines in children and adolescents. Among them, China (10, 43.5%) and United Stated (9, 39.1%) were the top two countries with the most trials. BNT162b2 was the most common vaccine, which is under testing.

**Conclusion:** Some of the COVID-19 vaccines have potential protective effects in children and adolescents, but awareness is needed to monitor possible adverse effects after injection. Clinical trials of the COVID-19 vaccine in children and adolescents with long follow-up, large sample size, and different vaccines are still urgently needed.

## Background

One and a half year has passed since the coronavirus disease 2019 (COVID-19) pandemic outbreak. Yet the epidemic is still not under better control. With over 200 million people already infected by severe acute respiratory syndrome coronavirus 2 (SARS-COV-2) and over 4 million deaths, COVID-19 has brought great suffering and devastation to people worldwide.

Vaccines, as an effective way to prevent and control disease infections, stimulate the human immune system to produce antibodies, thus increasing immunity to the disease and achieving protection for the immunized individual [1]. Vaccination aims to curb the spread of disease and improve the immunity of populations to achieve herd immunity. As of 26 July 2021, twenty-one COVID-19 vaccines have been approved worldwide, however, none of them are verified for children and adolescents. Given that children and adolescents account for approximately one quarter of the world’s population [2], promoting vaccination of children and adolescents is also one of the potential alternatives for the future to end the spread of COVID-19.

The development of the COVID-19 vaccine has been in full swing since the COVID-19 outbreak. Vaccines have also been shown to be effective and safe in many countries. Studies have shown that the current COVID-19 vaccine has good protection and safety in adults [3-6]. Several international organizations and countries have also developed guidelines for COVID-19 vaccination, including vaccination of special populations, management of adverse reactions, and cautions for vaccination [7-9]. However, the efficacy of protection and adverse effects of COVID-19 vaccine in children and adolescents remains unclear despite a large number of clinical trials being conducted. Furthermore, children and adolescents have less severe COVID-19 symptoms than adults [10], and children and adolescents play a minimal role in spreading the infection to others. Therefore, a large number of clinical studies are still needed to determine whether COVID-19 vaccination should be mandatory for children now [11]. In addition, children as a special group, the attitude of parents or guardians towards the COVID-19 vaccine is also an essential factor affecting children’s vaccination. Therefore, to explore and promote COVID-19 vaccination in children and adolescents, The National Clinical Research Center for Child Health and Disorders (Chongqing, China) initiated an international guideline for the management of COVID-19 in children and adolescents [12] that included the question of whether children and adolescents should be vaccinated with COVID-19. To answer this question, we propose to produce a systematic review to identify the safety, immunogenicity, and protective efficacy of the COVID-19 vaccine in children and adolescents. Meanwhile, we plan to include and analyze the clinical trials related to the COVDI-19 vaccine for children and adolescents that have already been registered to inform and advise on future studies.

## Methods

We conducted this systematic review in accordance with the Preferred Reporting Items for Systematic Reviews and Meta-Analysis (PRISMA) (**see supplementary 1 for PRISMA checklist**) [13]. In addition, the Cochrane Handbook for Systematic Reviews of Interventions was followed to develop this systematic review [14]. We have registered this systematic review at OSF REGISTRIES and the registration DOI is <u>10.17605/OSF.IO/JC32H</u>.

### 2.1 Inclusion and exclusion criteria

We included published research or ongoing clinical trials related to the safety, immunogenicity, and efficacy of COVID-19 vaccine in children or adolescents (aged ≤18 years). The study design was limited to primary studies, including randomized clinical trials (RCTs), non-RCTs or observational studies. We also include the ongoing trials registered at the International Clinical Trials Registry Platform (ICTRP).

We excluded studies fulfilling the following criteria: 1) could not extract data on children or adolescents; 2) could not access the full text; 3) conference proceedings; 4) study protocols. For ongoing trials, we will only include registration records where the study aims to determine the safety, immunogenicity or efficacy of COVID-19 vaccine in children and adolescents.

### 2.2 Search strategy

We systematically searched databases including Medline (via PubMed), Web of Science, World Health Organization (WHO) COVID-19 database, and China National Knowledge Infrastructure (CNKI), from their inception to 23 July 2021 to find out original studies related to the safety, immunogenicity, and efficacy of COVID-19 vaccine in children or adolescents. The search terms included “COVID-19”, “SARS-Cov-2”, “2019-nCov”, “Coronavirus disease 2019”, “adolescent”, “young”, “pediatrics”, “children”, “infant”, “newborn”, “neonates”, “youth”, and “vaccine” (**see detailed search strategy in supplementary 2**). All search strategies were developed and retrieved independently by two researchers (ML and XL) and then cross-checked. We first developed a search strategy for Medline after several attempts, and the search strategies for the other databases were then adapted from Medline. In addition, we also searched ICTRP to collect related ongoing trials. The search terms included “COVID-19”, “SARS-Cov-2”, adolescent”, “young”, “pediatrics”, “children” and “vaccine”. We also searched Google Scholar and reference lists of identified articles to avoid missing important literature.

### 2.3 Literature screening

The screening process included three phases. First, a reviewer removed duplicates of the retrieved literature. Following this, four reviewers (ML, XL, RL and QS) screened all identified records independently by reading titles and abstracts. And then if there was insufficient information based on the title and abstract, the full text was obtained for review. Disagreements were solved by consensus with the senior researcher (YC). We used Endnote 20.0.1 in the whole screening process.

### 2.4 Data extraction

The following data was extracted from the original studies: 1) basic information: publication date, country, study design, name of vaccine; 2) participants information: age, sample size, female/male number; and 3) outcome information: safety, immunogenicity, and efficacy of COVID-19. As for ongoing clinical trials, we extracted data including registration date, country, recruitment status, participants’ age, target sample size, intervention, and primary outcome. All the data were independently extracted by two reviewers (ML and XL) using a predesigned table.

### 2.5 Risk of bias assessment

Two reviewers (ML and XL) assessed the methodological quality of original studies to ensure the reliability of the included studies to support our findings. We used the Risk of Bias tool recommended by Cochrane Collaboration [15] to assess randomized trials with six domains of bias (selection bias, performance bias, detection bias, attrition bias, reporting bias, and other bias). We used the Newcastle-Ottawa Scale (NOS) to assess the quality of the included case-control and cohort studies (selection, comparability, and exposure) [16]. We used the checklist proposed by Murad *et al* [17] to assess case series and case reports (selection, ascertainment, causality and reporting) and used the checklist proposed by the Joanna Briggs Institute (JBI) with eleven items to assess cross-sectional study [18].

### 2.6 Data analysis

For quantitative analysis, meta-analysis was planned if two or more separate studies were included, and the heterogeneity was good. For dichotomous outcome data, the risk ratio (RR) or odds ratio (OR) was used to calculate the effect measures. For continuous outcome data, the mean difference (or difference in means) or standardized mean difference (SMD) was used. When the data are conveniently available as summary statistics from each intervention group, the inverse-variance method was implemented directly. Meanwhile, we planned to use both random-effects and fixed-effects methods to perform the meta-analysis. Besides, a statistical test for heterogeneity will be conducted and the Chi-squared (χ^2^) test will be included in the forest plots. The meta-analyses are planned to be performed in RevMan 5.4.1 (The Cochrane Collaboration, 2020.) and Stata/SE 15.1 (Copyright 1985-2017 StataCorp LLC) software.

For qualitative analysis, we planned to conduct structured methods with the topic of safety, immunogenicity, and efficacy of COVID-19 vaccine and ongoing clinical trials in children or adolescents. Microsoft Excel 16.51 (2019) was used for data processing and analysis. For ongoing clinical trials, we will analyze the status of ongoing trials of COVID-19 vaccines in children or adolescents in different countries and territories. Adobe Illustrator will be used to map ongoing clinical trials of COVID-19 vaccine in children or adolescents worldwide.

## Results

### 3.1 creening result

A total of 3092 records were identified, of which, 931 were excluded as duplicates. After screening the title and full text, finally, we included eight published studies [19-26] with 2851 children or adolescents and 28 ongoing clinical trials with 122,442 target participants. The study selection process was detailed in **Figure 1**.

**Figure 1.**
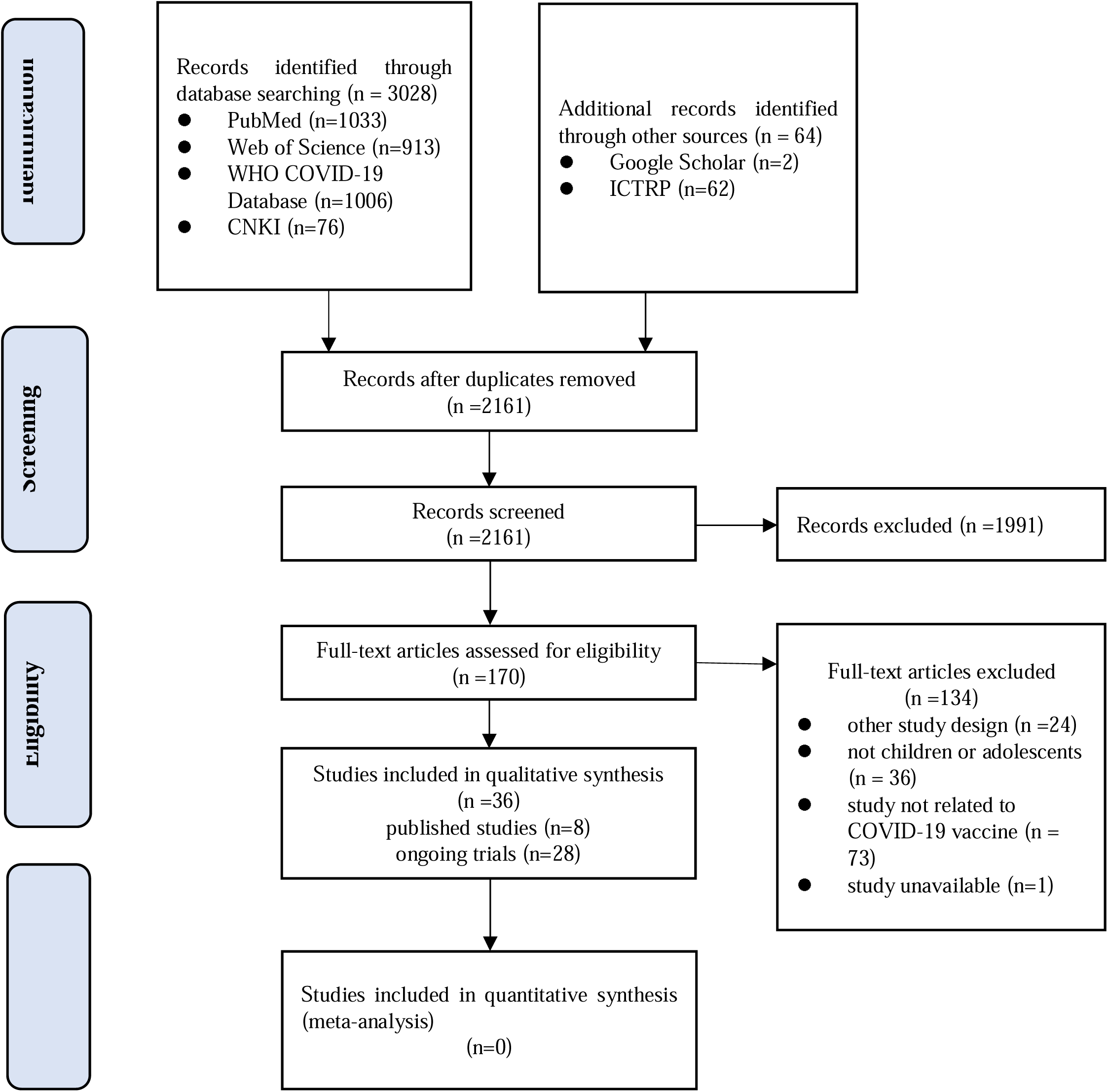
Study selection process. (WHO: World Health Organization; COVID-19: coronavirus disease 2019; CNKI: China National Knowledge Infrastructure; ICTRP: International Clinical Trials Registry Platform)

### 3.2 Characteristics of included clinical studies

Among eight published studies included, two (25.0%) were RCTs [19-20], two (25.0%) were case series [21-22] and four (50.0%) were case reports [23-26]. Five (62.5%) were conducted by the United States, and one each from China, France and Israel. Participants in the studies were between 12-18 years old, except for one study that included participants aged 3-17 years old. Except for participants in one study who received CoronaVac COVID-19 vaccine developed by Sinovac Life Sciences, participants in the other seven were received BNT162b2 mRNA COVID-19 vaccine developed by Pfizer-BioNTech. The characteristics of included studies were summarized in **Table 1**.

**Table 1.** Basic characteristics of included clinical studies (N=8)

| No | Country | Study design | Participants | Sample size | Name of vaccine | Follow-up duration | Funding |
| --- | --- | --- | --- | --- | --- | --- | --- |
| Han *et al*, 2021 | China | RCT Phase 1-2 | Healthy children and adolescents aged 3-17 years | 552 | CoronaVac | 4.1 months | Public/nonprofit (Chinese National Key Research and Development Program and Beijing Science and Technology Program) |
| Freneck *et al*, 2021 | USA | RCT Phase 3 | Adolescents aged 12-15 years with no previous Covid-19 diagnosis or SARS-CoV-2 infection | 2264 | BNT162b2 | 4.7 months | Private (BioNTech and Pfizer) |
| Riviere *et al*, 2021 | France | Case series | Adolescents and young adults aged 16 years with solid tumor older than | 11 | BNT162b2 | NR | NR |
| Snapiri *et al*, 2021 | Israel | Case series | Adolescents aged 16-18 years | 7 | BNT162b2 | NR | None |
| Minocha *et al*, 2021 | USA | Case report | An adolescent aged 17 years | 1 | BNT162b2 | 2 weeks | NR |
| McLean *et al*, | USA | Case report | A previously healthy | 1 | BNT162b2 | 2 weeks | NR |
| 2021 |  |  | adolescent<br>aged 16<br>years |  |  |  |  |
| Marshall<br>*et al*,<br>2021 | USA | Case<br>report | Healthy<br>adolescents<br>14-18 years | 6 | BNT162b<br>2 | unclear | None |
| Schauer<br>*et al*,<br>2021 | USA | Case<br>report | Children and<br>adolescents<br>aged 12-17<br>years | 13 | BNT162b<br>2 | 3 months | NR |

\*NR: not report; M: male; F: female

### 3.2 Quality of included studies

We assessed the methodological quality of included studies (**Table 2**). The overall quality of the two RCTs was high and had low risk of bias. As for the rest case series and case reports, we didn’t assess items of “was there a challenge/rechallenge phenomenon” and “Was there a dose-response effect?”, because of not application. One study fulfilled five items, three fulfilled four items, and one each fulfilled three and two items. The method of case selection for all case series and case reports is unclear. Only one-third of the case reports and case series reported items “were other alternative causes that may explain the observation ruled out?”, and half study follow-up was not long enough for outcomes to occur.

**Table 2.**
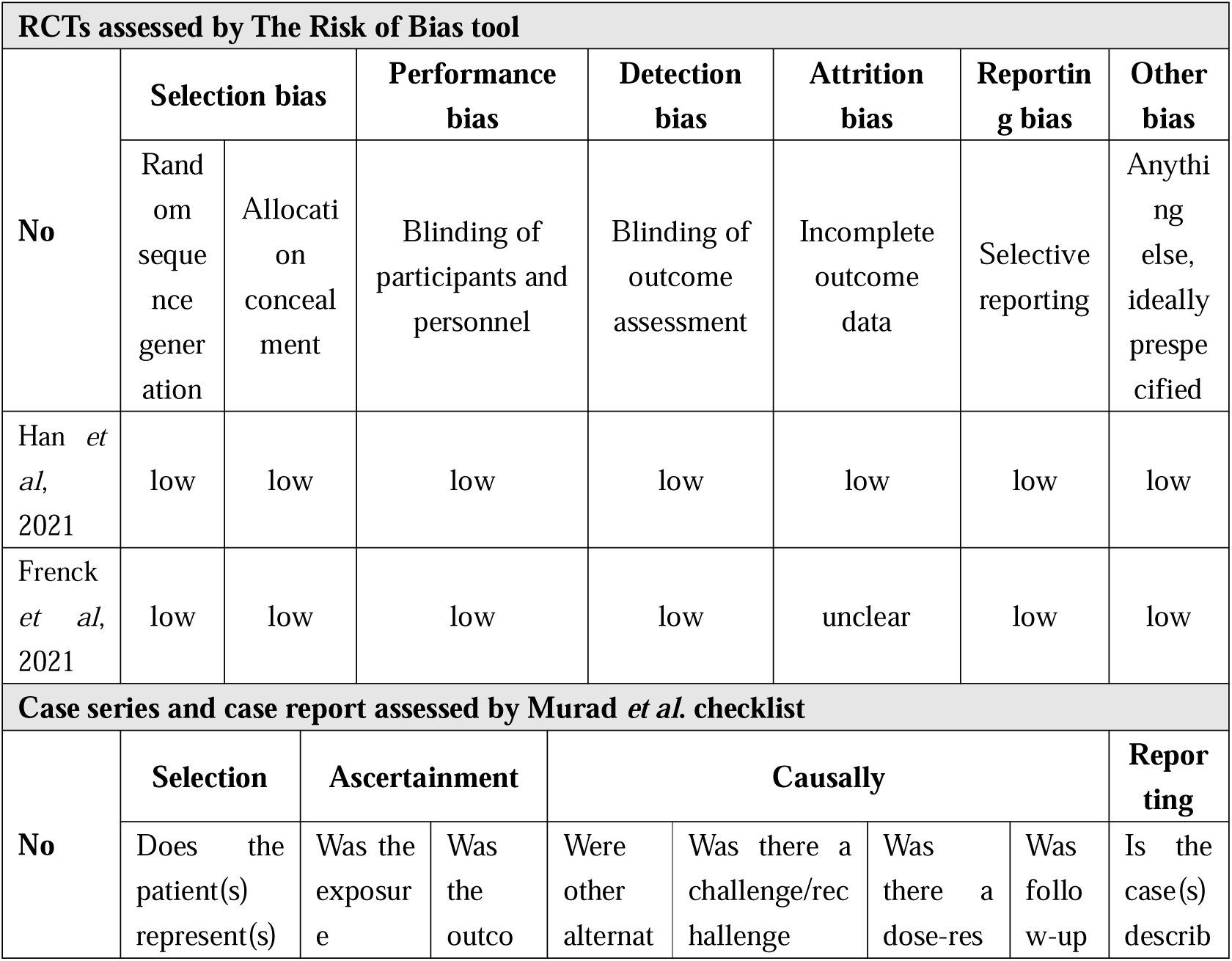
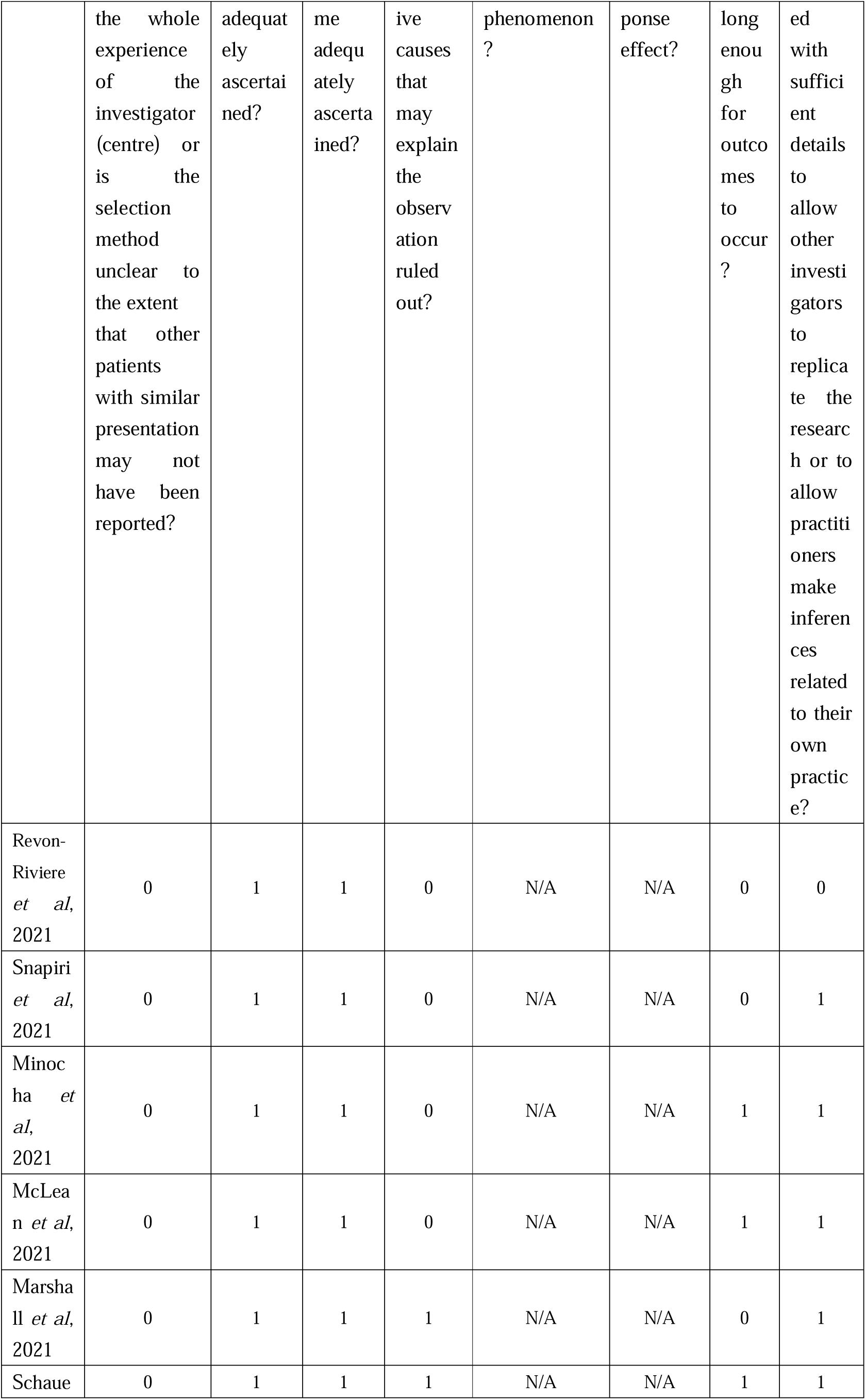
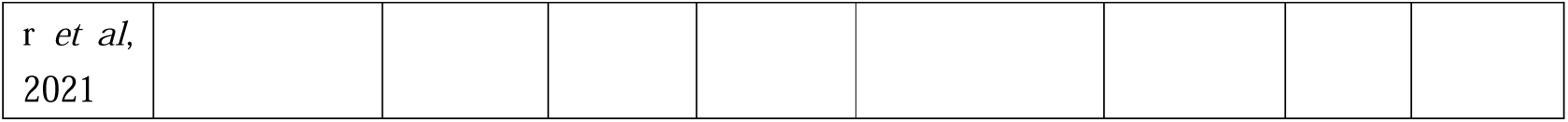
Quality assessment of included studies.

### 3.3 Safety of COVID-19 vaccine

Results of two RCTs [20-21] showed that after COVID-19 vaccination in healthy children and adolescents, the most common adverse event was injection site pain. Besides that, fever, headache, and fatigue were also frequently reported. Most adverse events were not severe. No deaths case was reported. With a similar result, a case series [22] included 13 patients with solid tumor showed that injection site pain is also the most frequent adverse event (6 patients), which is mild-to-moderate.

Besides, a few diagnosed myocarditis and/or pericarditis cases related to COVID-19 vaccine were reported in some studies. All cases occurred following the second dose of BNT162b mRNA COVID-19 vaccination. We summarized the basic information of 28 cases from included studies (**Table 3**). The median age was 15.8 years (range, 12-18 years). Most patients were male (27, 96.4%). Median days of onset after vaccination was 2.5 days (range, 1-4 days). All the patients had chest pain.

**Table 3.** Basic information of diagnosed myocarditis and/or pericarditis cases.

| Case | Vaccination | Diagnosis | Time of onset (days) | Symptoms | Length of hospitalization (days) |
| --- | --- | --- | --- | --- | --- |
| Case 1 [15] | BNT162b2, second dose | Perimyocarditis | 3 | Chest pain | 4 |
| Case 2 [15] | BNT162b2, second dose | Perimyocarditis | 1 | Chest pain | 6 |
| Case 3 [15] | BNT162b2, second dose | Perimyocarditis | 2 | Chest pain, cough | 6 |
| Case 4 [15] | BNT162b2, second dose | Perimyocarditis | 3 | Chest pain, nausea | 4 |
| Case 5 [15] | BNT162b2, second dose | Perimyocarditis | 1 | Chest pain, headache | 5 |
| Case 6 [15] | BNT162b2, second dose | Perimyocarditis | 2 | Chest pain, dyspnea, diarrhea, fever | 5 |
| Case 7 | BNT162b2, second dose | Perimyocarditis | 3 | Chest pain, dyspnea | 3 |
| [16] |  |  |  |  |  |
| Case 8 [17] | BNT162b2, second dose | Myocarditis | 1 | Chest pain, fever, body aches, | 6 |
| Case 9 [18] | BNT162b2, second dose | Myopericarditis | 2.5 | Chest pain | 6 |
| Case 10 [18] | BNT162b2, second dose | Myocarditis | 2 | Chest pain, bilateral arm pain, fever, fatigue, nausea, vomiting, anorexia, headache | 6 |
| Case 11 [18] | BNT162b2, second dose | Myopericarditis | 2 | Chest pain, bilateral arm pain | 2 |
| Case 12 [18] | BNT162b2, second dose | Myocarditis | 2 | Chest pain, fever, fatigue, nausea | 4 |
| Case 13 [18] | BNT162b2, second dose | Myocarditis | 4 | Chest pain, bilateral arm pain, fever, nausea, vomiting, anorexia, SOB, palpitations | 5 |
| Case 14 [18] | BNT162b2, second dose | Myocarditis | 3 | Chest pain, SOB | 3 |
| Case 15 [18] | BNT162b2, second dose | Myopericarditis | 2 | Chest pain, fever, SOB | 4 |
| Case 16 [19] | BNT162b2, second dose | Myopericarditis | 2 | Chest pain, fever, chills, myalgias, headache, SOB | 1 |
| Case 17 [19] | BNT162b2, second dose | Myopericarditis | 2 | Chest pain, fever, myalgias | 1 |
| Case 18 [19] | BNT162b2, second dose | Myopericarditis | 3 | Chest pain, myalgias, headache | 3 |
| Case 19 [19] | BNT162b2, second dose | Myopericarditis | 3 | Chest pain, fever, malaise | 1 |
| Case 20 [19] | BNT162b2, second dose | Myopericarditis | 2 | Chest pain, myalgias, SOB | 2 |
| Case 21 [19] | BNT162b2, second dose | Myopericarditis | 3 | Chest pain, vomiting | 1 |
| Case | BNT162b2, | Myopericarditis | 3 | Chest pain, fevers, SOB | 3 |
| 22<br>[19] | second dose |  |  |  |  |
| Case<br>23<br>[19] | BNT162b2,<br>second dose | Myopericarditis | 3 | Chest pain, chills | 3 |
| Case<br>24<br>[19] | BNT162b2,<br>second dose | Myopericarditis | 3 | Chest pain | 2 |
| Case<br>25<br>[19] | BNT162b2,<br>second dose | Myopericarditis | 3 | Chest pain, fever, headache | 3 |
| Case<br>26<br>[19] | BNT162b2,<br>second dose | Myopericarditis | 4 | Chest pain, malaise, SOB | 2 |
| Case<br>27<br>[19] | BNT162b2,<br>second dose | Myopericarditis | 2 | Chest pain, SOB | 2 |
| Case<br>28<br>[19] | BNT162b2,<br>second dose | Myopericarditis | 3 | Chest pain | 2 |

M: male; F: female; SOB: shortness of breath

### 3.4 Immunogenicity of COVID-19 vaccine

Two RCTs indicated that COVID-19 vaccines (CoronaVac and BNT162b2) were immunogenic in children and adolescents. Frenck *et al*. [20] reported that the immune response to BNT162b2 in 12-15 years old adolescents was noninferior to that in young adults (geometric mean ratio (GMR)=1.75, 95%CI: 1.47∼2.10), which even indicated a greater response in 12-15 years group. Han *et al*. [19] found that in Phase 1, the seroconversion of neutralizing antibody after the second dose was 100% both in 1.5μg group and 3.0μg group with GMT of 55·0 (95% CI 38.9–77.9) and 117·4 (87.8–157.0), respectively (*P*=0.0012). In Phase 2, the seroconversion was 96·8% (95%CI: 93·1–98·8) and 100% (95%CI: 98.0-100.0) in 1.5μg group and 3.0μg group (*P*=0.030).

### 3.5 Efficacy of COVID-19 vaccine

There is only one RCT [20] that showed that the efficacy of the BNY162b2 vaccine in children and adolescents is 100% (95%CI: 75.3∼100).

### 3.6 Ongoing clinical studies

We identified 28 ongoing eligible clinical trials with 122,442 target sample size (**see Supplementary 3**). Twenty-three are interventional studies (including one Phase 1 trial; six Phase1/2 trials; six Phase 2 trials; four Phase 2/3 trials; three Phase 3 trials; one Phase 4 trial; and one not applicable) and five observational studies. The minimum age of participants is 6 months (NCT04816643, NCT04796896, NCT04299724, NCT04276896). Twenty-seven trials planned to use 15 different vaccine candidates as intervention with five types, including RNA (13 trials), inactivated (7 trials), protein subunit (4 trials), non replicating viral vector (4 trials), and replicating viral vector (1 trial).

Fifteen countries are conducting interventional clinical trials of COVID-19 vaccines in children and adolescents. Among them, China (10, 43.5%) and United Stated (9, 39.1%) were the top two countries with the most trials. BNT162b2 was the most common vaccine, which is under testing. Figure 2 showed countries with ongoing clinical trials and vaccines used in trials.

**Figure 2.**
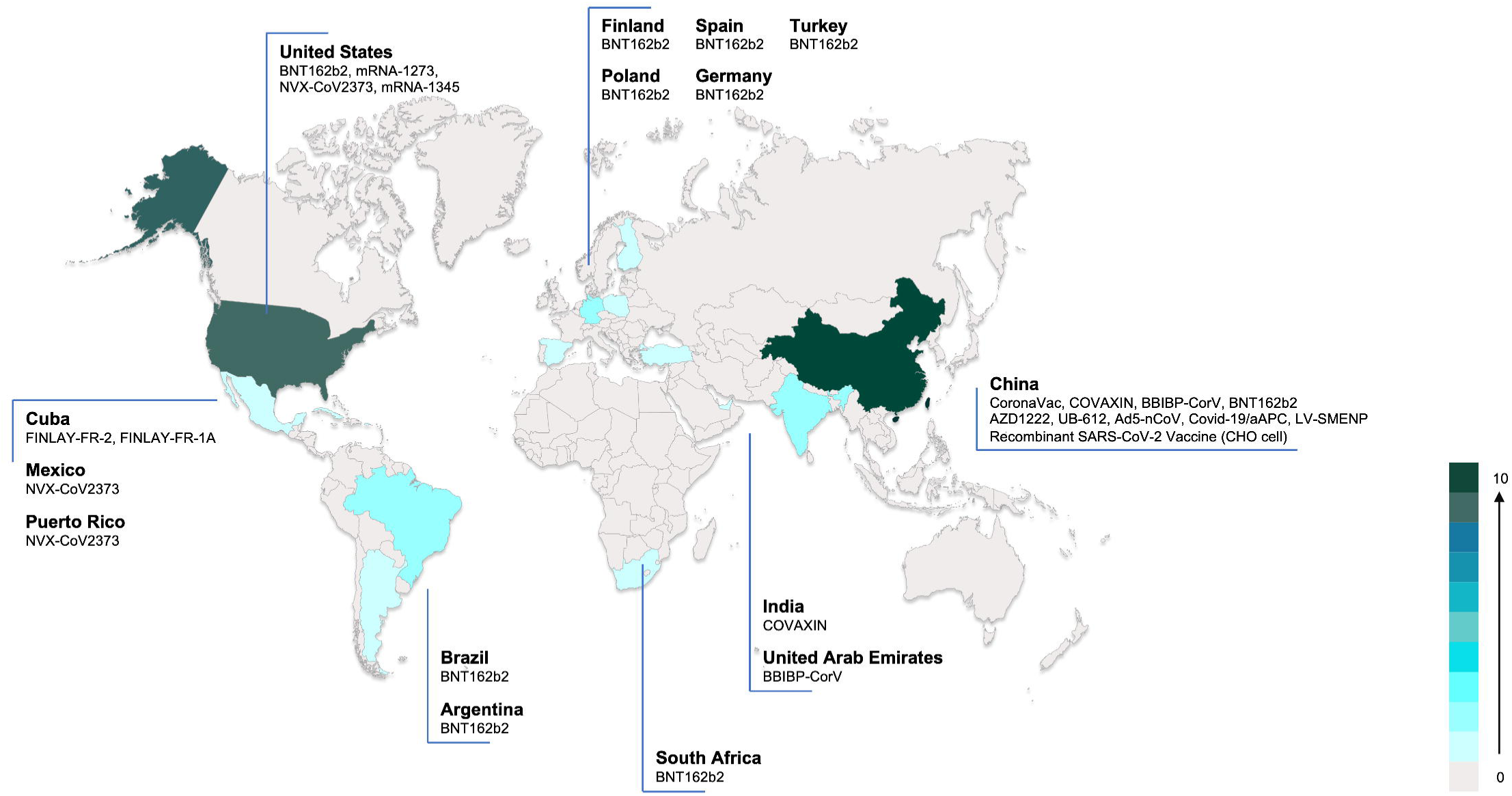
ongoing COVID-19 vaccine trials in children and adolescents worldwide (Notes: only interventional trials registered at ICTRP were shown in the figure. Color in the figure indicated that the number of ongoing vaccine trials in children and adolescents per country.)

## Discussion

### Principal findings

Our review included eight original studies and 28 clinical trial registries of COVID-19 vaccine in children and adolescents. The results showed that selected COVID-19 vaccines had a good safety profile in children and adolescents, with mostly mild and moderate adverse effects, mostly including injection site pain, fatigue, headache, and chest pain. Besides, some studies reported a few cases of myocarditis and pericarditis. Regarding immunogenicity, two RCTs showed that the immune response to BNT162b2 in adolescents aged 12-15 years was non-inferior to that in young people aged 16-25 years, while a stronger immune response was detected with 3μg CoronaVac injection. Only one single RCT showed no cases of infection in 12-15 years received BNT162b2, with the efficacy of 100% (95% CI: 75.3 to 100). Clinical trials on children and adolescents are conducting all over the world with various vaccines.

Children and adolescents, as a special population, have many influencing factors to consider when administering vaccines, with vaccine efficacy and safety being the most important considerations for children and their parents [27]. It is therefore important to demonstrate that vaccines are safe and protective before they are administered to children and adolescents. Earlier in 2009 during the influenza A (H1N1) outbreak, approximately 9.8% of children developed symptoms [28] and when the vaccine was administered, approximately 90.3% of children and adolescents aged 10-17 years developed protective antibodies, and no serious adverse reactions were seen [29-30]. Similarly, when the COVID-19 outbreak emerged, researchers actively promoted the development of a vaccine with the expectation that herd immunity could be reached after vaccination. Our study showed that some vaccines have now been developed in pediatric populations with associated RCTs and proven protective efficacy and safety, however, these two studies both have limitations on small sample size and lack of long-term safety and immunogenicity data, for example, myocarditis and pericarditis should be closely monitored. Most cases of myocarditis and pericarditis associated with the COVID-19 vaccine were mild and mostly in males. According to Washington State Department of Health data on immunization, Schauer *et al*. [26] estimated a possible incidence of 0.008% in adolescents 16-17 years of age and 0.01% in those 12 through 15 years of age following the second dose.

Another population to consider for vaccination of children and adolescents is multisystemic inflammatory syndrome (MIS). In April 2020, children with presentations similar to incomplete Kawasaki disease (KD) or toxic shock syndrome were documented in reports from the UK [31]. Since then, such children have been reported in other parts of the world [32-34]. This is termed multisystemic inflammatory syndrome in children (MIS-C). Study showed that the pooled proportions of MIS-C in Hispanic and Black cases were 34.6% (95% CI, 28.3–40.9) and 31.5% (95% CI, 24.8–38.1), respectively [35]. The overall mortality of MIS-C is approximately 1–2% [36]. Our review did not involve the children with MIS-C. Therefore, it is unclear whether children with SARS-CoV-2 infection complicated by multisystemic inflammatory syndrome should be vaccinated with COVID-19. The decision to vaccinate should be made by weighing the risk of exposure, reinfection and severe disease following infection against the uncertain safety of vaccination in such individuals. Whereas no directly relevant studies have confirmed the association of MIS-C with COVID-19 vaccination, a systematic review published in 2017 [37] identified 27 observational studies and case reports of KD showed that diphtheria-tetanus-pertussis (DTP)-containing vaccines, Haemophilus influenzae type b (Hib) conjugate vaccine, influenza vaccine, hepatitis B vaccine, 4-component meningococcal serogroup B (4CMenB) vaccine, measles-mumps-rubella (MMR)/MMR-varicella vaccines, pneumococcal conjugate vaccine (PCV), rotavirus vaccine (RV), yellow fever vaccine, and Japanese encephalitis vaccine did not increase the risk of KD following any of the above immunizations. Thus, children and adolescents at high risk of severe COVID-19 or those with specific comorbidities should be considered a priority of vaccination, and more research is needed to clarify whether the COVID-19 vaccine can decrease the risks and bring benefits.

To date, there are 21 approved COVID-19 vaccines worldwide, more than 1/3 of which are inactivated, and 138 vaccines in development and exploitation, while more than 300 clinical trials of COVID-19 vaccines have been registered or published [38-39]. Studies have shown that most COVID-19 vaccines are safe and effective in adults (>18y). Overall, in phase 2/3 RCTs, mRNA- and adenoviral vector-based COVID-19 vaccines had 94.6% (95% CI 0.936-0.954) and 80.2% (95% CI 0.56-0.93) efficacy, respectively [3-5], with good acceptability [6] and safety [40]. However, there are only two RCTs published in peer reviewed journals and found that BNT162b2 and CoronaVac are safe and effective in children and adolescents. Institutions including WHO, Centers for Disease Control and Prevention (CDC), the Food and Drug Administration (FDA), the Canadian Paediatric Society authorized emergency use of Pfizer-BioNTech COVID-19 Vaccine in children and adolescents [41-44]. European Medicines Agency (EMA) recommended children aged 12 to 17 years are able to use Spikevax (previously COVID-19 Vaccine Moderna) vaccine according to an ongoing study [45]. At present, vaccines recommended in the guidelines are for countries that have made vaccine progress among children or adolescents. We still need to wait for more evidence from ongoing trials for some low- and middle-income countries with vaccine shortages. With the conduction of more than twenty clinical trials, their findings may continue to offer clues of better protecting younger generations away from COVID-19.

However, public health authorities in countries that approved COVID-19 vaccine in children and adolescents should also take more considerations into decision making. European Centre for Disease Prevention and Control proposed eight considerations on the overall potential public health impact [46]. Opel et al. suggested nine criteria to consider when evaluating antigens for inclusion in mandatory school immunization programs. Also, four additional criteria need to be met for mandatory COVID-19 vaccination of children, but we currently know too little about the performance of any COVID-19 vaccine or the epidemiology of SARS-CoV-2 in children to make any definitive judgment about whether COVID-19 vaccine should be mandatory in children [11]. Authorities could closely monitor and continually assess the benefits and potential risks of vaccination in children and adolescents. In addition, the acceptability of the COVID-19 vaccine to children, as well as to parents, is a major influencing factor on whether children can be vaccinated. Studies have shown that approximately 80% of parents are reluctant to enroll their children in clinical studies of the COVID-19 vaccine [47] and approximately half of children are unwilling to take the COVID-19 vaccine [48]. Therefore, it is necessary to educate parents and children about the vaccine to increase vaccination rates while ensuring its efficacy and safety [49]. Furthermore, factors such as national policy, religion, culture, and other routine immunization procedures need to be taken into account for the administration of COVID-19 vaccine to children.

### Potential impact for future research and practice

While our study included only two RCTs on COVID-19 vaccination in children and adolescents, which contained CoronaVac developed by Sinovac and BNT162b2 developed by Pfizer/BioNTech, as yet the vast majority of vaccines have not been conducted or are being conducted with COVID-19 vaccine in clinical studies in children and adolescent populations. For future research, we recommend the following three aspects. First, we should continue to conduct clinical studies on the protective efficacy and safety of COVID-19 vaccine in children and adolescents; second, we should conduct systematic reviews of factors affecting COVID-19 vaccination in children and adolescents, willingness to vaccinate, and methods to promote vaccination. In addition, we should update this systematic review when enough studies of COVID-19 in children and adolescents, especially RCTs, are available; third, we should develop evidence-based guidelines for COVID-19 vaccination in children and adolescents to promote and standardize vaccination in children and adolescents. Policymakers should develop policies for COVID-19 vaccination in children and adolescents based on the best current evidence in the future, and parents should be guided by policies that actively encourage and support their children to be vaccinated against COVID-19.

### Strengths and limitations

This paper is, to the best of our knowledge, the first systematic review of the safety, immunogenicity, and protective efficacy of COVID-19 vaccination in children and adolescents. We systematically searched key databases as well as websites to conduct a comprehensive evaluation and analysis of published studies and registry data records. However, this paper also has some limitations. First, we didn’t conduct a meta-analysis in this study, because of the great heterogeneity among participants, outcomes, and study design. Second, this study only included articles published in English. However, as the limited evidence published, it is reasonable to expect that included studies up until this time covered most of the available knowledge. Finally, a few studies that included children and adolescents didn’t specify the age and outcome related to children and adolescents. Given the limitation of time, we excluded these studies and didn’t contact authors to access original data.

## Conclusion

Some of the COVID-19 vaccines have potential protective effects in children and adolescents, but awareness is needed to monitor their possible adverse effects after injection, especially myocarditis and pericarditis. Meanwhile, more clinical studies with long follow-up, large sample sizes, and different vaccine categories on COVID-19 in children and adolescents should be conducted in the future, and relevant evidence-based guidelines should be developed to inform policymakers, parents, and children and adolescents.

## Data Availability

The datasets used during the current study are available from the corresponding author on reasonable request.

## Author contributions

**Conceptualization**: Yaolong Chen, Enmei Liu and Qiu Li. **Study design:** Yaolong Chen and Qiu Li. **Literature search**: Meng Lv and Xufei Luo. **Figures and Table**: Meng Lv and Xiao Liu. **Data collection**: Ruobing Lei, Quan Shen, Xufei Luo and Meng Lv. **Data analysis**: Meng Lv and Xufei Luo. **Data interpretation**: Yaolong Chen, Qiu Li, Enmei Liu and Meng Lv. **Writing**: Meng Lv, Xufei Luo and Yaolong Chen. **Supervision**: Yaolong Chen, Enmei Liu and Qiu Li. All authors provided input regarding the direction of the study and the content of the paper. All authors approved the final version of the paper.

## References

[1] CDC. Vaccines: The Basics. https://www.cdc.gov/vaccines/vpd/vpd-vac-basics.html

[2] The WORLD Bank. Population ages 0-14 (% of total population). https://data.worldbank.org/indicator/SP.POP.0014.TO.ZS

[3] Xing K, Tu XY, Liu M, et al. Efficacy and safety of COVID-19 vaccines: a systematic review. Zhongguo Dang Dai Er Ke Za Zhi. 2021;23(3):221–228. doi:10.7499/j.issn.1008-8830.2101133.

[4] Pormohammad A, Zarei M, Ghorbani S, et al. Efficacy and Safety of COVID-19 Vaccines: A Systematic Review and Meta-Analysis of Randomized Clinical Trials. Vaccines (Basel). 2021;9(5):467. Published 2021 May 6. doi:10.3390/vaccines9050467

[5] McDonald I, Murray SM, Reynolds CJ, et al. Comparative systematic review and meta-analysis of reactogenicity, immunogenicity and efficacy of vaccines against SARS-CoV-2. NPJ Vaccines. 2021;6(1):74. Published 2021 May 13. doi:10.1038/s41541-021-00336-1

[6] Wang Q, Yang L, Jin H, et al. Vaccination against COVID-A systematic review and meta-analysis of acceptability and its predictors [published online ahead of print, 2021 Jun 22]. Prev Med. 2021;150:106694. doi:10.1016/j.ypmed.2021.106694

[7] Pfaar O, Klimek L, Hamelmann E, et al. COVID-19 vaccination of patients with allergies and type-2 inflammation with concurrent antibody therapy (biologicals) - A Position Paper of the German Society of Allergology and Clinical Immunology (DGAKI) and the German Society for Applied Allergology (AeDA). Allergol Select. 2021;5:140–147. Published 2021 Apr 1. doi:10.5414/ALX02241E

[8] Hazlewood GS, Pardo JP, Barnabe C, et al. Canadian Rheumatology Association Recommendation for the Use of COVID-19 Vaccination for Patients With Autoimmune Rheumatic Diseases. J Rheumatol. 2021;48(8):1330–1339. doi:10.3899/jrheum.210288

[9] WHO. Interim recommendations for use of the inactivated COVID-19 vaccine, CoronaVac, developed by Sinovac. 2021. https://www.who.int/publications/i/item/WHO-2019-nCoV-vaccines-SAGE_recommendation-Sinovac-CoronaVac-2021.1

[10] Zimmermann P, Curtis N. Why is COVID-19 less severe in children? A review of the proposed mechanisms underlying the age-related difference in severity of SARS-CoV-2 infectionsArchives of Disease in Childhood 2021;106:429–439.

[11] Opel DJ, Diekema DS, Ross LF. Should We Mandate a COVID-19 Vaccine for Children?. JAMA Pediatr. 2021;175(2):125–126. doi:10.1001/jamapediatrics.2020.3019

[12] Zhou Q, Li W, Zhao S, et al. Guidelines for the Management of Children and Adolescent with COVID-19: protocol for an update. Transl Pediatr. 2021;10(1):177–182. doi:10.21037/tp-20-290

[13] Moher D, Liberati A, Tetzlaff J, et al. Preferred reporting items for systematic reviews and meta-analyses: the PRISMA statement[J]. Int J Surg,2010, 8(5): 336–341.

[14] Higgins J, Thomas J. Cochrane Handbook for Systematic Reviews of Interventions. 2021. https://training.cochrane.org/handbook/current.

[15] Higgins J P T, Altman D G, Gøtzsche P C, et al. The Cochrane Collaboration’s tool for assessing risk of bias in randomised trials[J]. BMJ, 2011, 343.

[16] Peterson J, Welch V, Losos M, et al. The Newcastle-Ottawa scale (NOS) for assessing the quality of nonrandomised studies in meta-analyses[J]. Ottawa: Ottawa Hospital Research Institute, 2011: 1–12.

[17] Murad M H, Sultan S, Haffar S, et al. Methodological quality and synthesis of case series and case reports[J]. BMJ evidence-based medicine, 2018, 23(2): 60–63.

[18] Joanna Briggs Institute, Joanna Briggs Institute. Checklist for analytical cross sectional studies[J]. Adelaide: The Joanna Briggs Institute, 2017.

[19] Han B, Song Y, Li C, et al. Safety, tolerability, and immunogenicity of an inactivated SARS-CoV-2 vaccine (CoronaVac) in healthy children and adolescents: a double-blind, randomised, controlled, phase 1/2 clinical trial[J]. The Lancet Infectious Diseases, 2021.

[20] Frenck Jr R W, Klein N P, Kitchin N, et al. Safety, immunogenicity, and efficacy of the BNT162b2 Covid-19 vaccine in adolescents[J]. New England Journal of Medicine, 2021.

[21] Revon-Riviere G, Ninove L, Min V, et al. BNT162b2 mRNA Covid-19 Vaccine in AYA with cancer: a monocentric experience[J]. European Journal of Cancer, 2021.

[22] Snapiri O, Shirman N, Weissbach A, et al. Transient Cardiac Injury in Adolescents Receiving the BNT162b2 mRNA COVID-19 Vaccine[J]. The Pediatric Infectious Disease Journal, 2021.

[23] Minocha P K, Better D, Singh R K, et al. Recurrence of Acute Myocarditis Temporally Associated With Receipt of the mRNA COVID-19 Vaccine in an Adolescent Male[J]. The Journal of Pediatrics, 2021.

[24] McLean K, Johnson T J. Myopericarditis in a Previously Healthy Adolescent Male Following COVIDC19 Vaccination: A Case Report[J]. Academic Emergency Medicine, 2021.

[25] Marshall M, Ferguson I D, Lewis P, et al. Symptomatic acute myocarditis in seven adolescents following Pfizer-BioNTech COVID□19 vaccination[J]. Pediatrics, 2021: 2.

[26] Schauer J, Buddhe S, Colyer J, et al. Myopericarditis after the Pfizer mRNA COVID-19 Vaccine in Adolescents[J]. The Journal of Pediatrics, 2021.

[27] Smith LE, Amlôt R, Weinman J, Yiend J, Rubin GJ. A systematic review of factors affecting vaccine uptake in young children. Vaccine. 2017;35(45):6059–6069. doi:10.1016/j.vaccine.2017.09.046

[28] Ruf BR, Knuf M. The burden of seasonal and pandemic influenza in infants and children. Eur J Pediatr. 2014;173(3):265–276. doi:10.1007/s00431-013-2023-6

[29] Wijnans L, de Bie S, Dieleman J, Bonhoeffer J, Sturkenboom M. Safety of pandemic H1N1 vaccines in children and adolescents. Vaccine. 2011;29(43):7559–7571. doi:10.1016/j.vaccine.2011.08.016

[30] Lu CY, Shao PL, Chang LY, et al. Immunogenicity and safety of a monovalent vaccine for the 2009 pandemic influenza virus A (H1N1) in children and adolescents. Vaccine. 2010;28(36):5864–5870. doi:10.1016/j.vaccine.2010.06.059

[31] Riphagen S, Gomez X, Gonzalez-Martinez C, et al. Hyperinflammatory shock in children during COVID-19 pandemic. Lancet 2020; 395:1607.

[32] Licciardi F, Pruccoli G, Denina M, et al. SARS-CoV-2-Induced Kawasaki-Like Hyperinflammatory Syndrome: A Novel COVID Phenotype in Children. Pediatrics 2020; 146.

[33] Verdoni L, Mazza A, Gervasoni A, et al. An outbreak of severe Kawasaki-like disease at the Italian epicentre of the SARS-CoV-2 epidemic: an observational cohort study. Lancet 2020; 395:1771.

[34] Whittaker E, Bamford A, Kenny J, et al. Clinical Characteristics of 58 Children With a Pediatric Inflammatory Multisystem Syndrome Temporally Associated With SARS-CoV-2. JAMA 2020; 324:259.

[35] Yasuhara J, Watanabe K, Takagi H, et al. COVID-19 and multisystem inflammatory syndrome in children: A systematic review and meta-analysis. Pediatr Pulmonol. 2021;56(5):837–848. doi:10.1002/ppul.25245

[36] Kaushik A, Gupta S, Sood M, et al. A Systematic Review of Multisystem Inflammatory Syndrome in Children Associated With SARS-CoV-2 Infection. Pediatr Infect Dis J. 2020;39(11):e340–e346. doi:10.1097/INF.0000000000002888

[37] Phuong L.K. Kawasaki disease and immunisation: a systematic review. Vaccine. 2017;35(14):1770–1779.

[38] VACCINES CANDIDATES BY TRIAL PHASE. https://covid19.trackvaccines.org/vaccines.

[39] COVID-19 NMA. https://covid-nma.com/vaccines/mapping/

[40] Kaur RJ, Dutta S, Bhardwaj P, et al. Adverse Events Reported From COVID-19 Vaccine Trials: A Systematic Review [published online ahead of print, 2021 Mar 27]. Indian J Clin Biochem. 2021;1–13. doi:10.1007/s12291-021-00968-z

[41] World Health Organization. Interim recommendations for use of the Pfizer–BioNTech COVID-19 vaccine, BNT162b2, under emergency use listing: interim guidance, first issued 8 January 2021, updated 15 June 2021[R]. World Health Organization, 2021.

[42] Centers for Disease Control and Prevention. COVID-19 Vaccines for Children and Teens. Updated July 23, 2021. https://www.cdc.gov/coronavirus/2019-ncov/vaccines/recommendations/adolescents.html

[43] Food and Drug Administration. Pfizer-BioNTech COVID-19 vaccine emergency use authorization. Silver Spring, MD: US Department of Health and Human Services, Food and Drug Administration; 2021. https://www.fda.gov/emergency-preparedness-and-response/coronavirus-disease-2019-covid-19/pfizer-biontech-covid-19-vaccine

[44] The Canadian Paediatric Society. COVID-19 vaccine for children. Updated July 12, 2021. https://www.cps.ca/en/documents/position/covid-19-vaccine-for-children

[45] European Medicines Agency. COVID-19 vaccine Spikevax approved for children aged 12 to 17 in EU. 23/07/2021. https://www.ema.europa.eu/en/news/covid-19-vaccine-spikevax-approved-children-aged-12-17-eu

[46] European Center for Disease Prevention and Control. Interim public health considerations for COVID-19 vaccination of adolescents in the EU/EEA. 1 June 2021. Stockholm: ECDC; 2021. https://www.ecdc.europa.eu/en/publications-data/interim-public-health-considerations-covid-19-vaccination-adolescents-eueea#no-link

[47] Goldman RD, Staubli G, Cotanda CP, et al. Factors associated with parents’ willingness to enroll their children in trials for COVID-19 vaccination. Hum Vaccin Immunother. 2021;17(6):1607–1611. doi:10.1080/21645515.2020.1834325

[48] Wang Q, Xiu S, Zhao S, et al. Vaccine Hesitancy: COVID-19 and Influenza Vaccine Willingness among Parents in Wuxi, China-A Cross-Sectional Study. Vaccines (Basel). 2021;9(4):342. Published 2021 Apr 1. doi:10.3390/vaccines9040342

[49] Nour R. A Systematic Review of Methods to Improve Attitudes Towards Childhood Vaccinations. Cureus. 2019;11(7):e5067. Published 2019 Jul 2. doi:10.7759/cureus.5067

