## supplemental file 1 for "Safety, Immunogenicity, and Efficacy of COVID-19 Vaccine in Children and Adolescents: A Systematic Review"

**Supplementary 1. Search Strategy**

**PubMed（N=1033）**

#1	"COVID-19"[Mesh]

#2	"SARS-CoV-2"[Mesh]

#3	"COVID-19"[Title/Abstract]

#4	"SARS-COV-2"[Title/Abstract]

#5	"SARS COV 2"[Title/Abstract]

#6	"Novel coronavirus"[Title/Abstract]

#7	"2019-novel coronavirus"[Title/Abstract]

#8	"Coronavirus disease-19"[Title/Abstract]

#9	"Coronavirus disease 19"[Title/Abstract]

#10	"Coronavirus disease 2019"[Title/Abstract]

#11	"COVID 19"[Title/Abstract]

#12	"Novel CoV"[Title/Abstract]

#13	"2019-nCoV"[Title/Abstract]

#14	"2019 nCoV"[Title/Abstract]

#15	"2019-CoV"[Title/Abstract]

#16	OR/#1-#15

#17	"vaccin*"[Title/Abstract]

#18	"Vaccines"[Mesh]

#19	#17 OR #18

#20	#16 AND #19

#21	"COVID-19 Vaccines"[Mesh]

#22	#20 OR #21

#23	"Adolescent"[Mesh]

#24	"Adolescen*"[Title/Abstract]

#25	"Teen*"[Title/Abstract]

#26	"Youth*"[Title/Abstract]

#27	"juvenile*"[Title/Abstract]

#28	"puberty"[Title/Abstract]

#29	"young*"[Title/Abstract]

#30	"Child" [Mesh]

#31	"child*" [Title/Abstract]

#32	"Pediatrics" [Mesh]

**Web of Science（N=913）**

#1	"SARS-CoV-2"[Topic]

#2	"Novel coronavirus"[Topic]

#3	"2019-novel coronavirus"[Topic]

#4	"Coronavirus disease 19"[Topic]

#5	"Coronavirus disease 2019"[Topic]

#6	"COVID-19"[Topic]

#7	"Novel CoV"[Topic]

#8	"2019-nCoV"[Topic]

#9	OR/#1-#8

#10	Adolescen* (Topic)

#11	young* (Topic)

#12	Pediatrics (Topic)

#13	Child* (Topic)

#14	Infant (Topic)

#15	newborn* (Topic)

#16	neonat* (Topic)

#17	Youth* (Topic)

#18	student* (Topic)

#19	OR/#10-#18

#20	vaccin* (Topic)

#21	#9 AND #19 AND 20

#22	#21 AND limit Database: Pediatrics (Research Areas)

**WHO（N=1006）**

#1	tw:(adolescen*)

#2	tw:(teen*)

#3	tw:(youth*)

#4	tw:(juvenile*)

#5	tw:(puberty)

#6	tw:(young*)

#7	tw:(child*)

#8	tw:(pediatric*)

#9	tw:(paediatric*)

#10	tw:(infant*)

#11	tw:(neonat*)

#12	tw:(newborn*)

#13	tw:(baby)

#14	tw:(trottie*)

#15	tw:(babies)

#16	tw:(kids)

#17	tw:(toddler*)

#18	tw:(pre-school*)

#19	tw:(preschool*)

#20	tw:(kindergarten*)

#21	tw:(kinder-garten*)

#22	tw:(girl*)

#23	tw:(boy*)

#24	tw:(student*)

#25	tw:(junior*)

#26	tw:(pubescent)

#27	OR/#1-#26

#28	tw:(vaccin**)

#29	#27 AND #28

#30	#29 AND la:("en")

**CNKI（N=76）**

#1	'儿童' (TOPIC)

#2	'幼儿' (TOPIC)

#3	'婴儿' (TOPIC)

#4	'新生儿' (TOPIC)

#5	'青少年' (TOPIC)

#6	'小儿' (TOPIC)

#7	OR/#1-#6

#8	'新型冠状病毒' (TOPIC)

#9	'COVID-19' (TOPIC)

#10	'COVID 19' (TOPIC)

#11	'2019-nCoV' (TOPIC)

#12	'2019 nCoV' (TOPIC)

#13	'2019-CoV' (TOPIC)

#14	'2019 CoV' (TOPIC)

#15	'SARS-CoV-2' (TOPIC)

#16	'SARS COV 2' (TOPIC)

#17	'新冠肺炎' (TOPIC)

#18	OR/#8-#17

#19	'疫苗' (TOPIC)

#20	'接种' (TOPIC)

#21	'免疫' (TOPIC)

#22	OR/#19-#21

#23	#1 AND #7 AND #18
