## supplemental table 2 for "Safety, Immunogenicity, and Efficacy of COVID-19 Vaccine in Children and Adolescents: A Systematic Review"

**Supplementary 2**

**Table1 Ongoing clinical trials on COVID-19 vaccination in children and adolescents**

| **Trial ID** | **Registration date** | **Recruitment stage** | **Country** | **Participants’ age** | **Target size** | **Study type** | **Intervention** | **Primary outcome** |
| --- | --- | --- | --- | --- | --- | --- | --- | --- |
| RPCEC00000374 | 10/06/2021 | Not Recruiting | Cuba | 3-18 | 350 | Interventional  Phase 1/2 | FINLAY-FR-2+FINLAY-FR-1A | Phase I: SAEs (It will measure as: -Occurrence of the SAE (Yes, No), - Duration (Time from start date until end date of event), -Description of the event, Result (Recovered, Recovered with squeals, Persists, Death, Unknown), - Causality (Causal association consistent with vaccination, Undetermined, Inconsistent causal association with vaccination, not classifiable). Measurement time: daily for 28 days after each dose.  Phase II: Concentration of specific anti-RBD IgG antibodies (Percentage of subjects with seroconversion 4-fold to pre-vaccination). Measurement time: Day 0, 42 and 70 |
| NCT04918797 | 18/05/2021 | Recruiting | India | 2-18 | 525 | Interventional  Phase 2/3 | COVAXIN | Reactogenicity; Immunogenicity |
| NCT04917523 | 04/06/2021 | Not recruiting | United Arab Emirates | ≥3 | 1800 | Interventional  Phase 3 | SARS-CoV-2 Vaccine (Vero Cell), Inactivated | The four-fold increase rate of anti-SARS-CoV-2 neutralizing antibody; GMT of anti-SARS-CoV-2 neutralizing antibody |
| NCT04916886 | 30/05/2021 | Recruiting | China | 6-59 | 2016 | Interventional  N/A | Recombinant Novel Coronavirus Vaccine (Adenovirus Type 5 Vector) | GMT of anti-SARS-CoV-2 specific neutralizing antibody |
| NCT04895982 | 05/05/2021 | Not recruiting | United States, Brazil, Germany | ≥2 | 360 | Interventional  Phase 2 | BNT162b2 | Percentage of participants reporting local reactions; Percentage of participants reporting systemic events; Percentage of participants reporting AEs; Percentage of participants reporting SAEs; GMTs of all participants, measured by SARS-CoV-2 neutralising titers, without serological or virological evidence of past SARS-CoV-2 infection and with an immunocompromised state, as specified in the protocol; Percentage of participants reporting AEs |
| NCT04894448 | 18/05/2021 | Recruiting | Canada | ≥16 | 500 | Observational | COVID-19 Vaccine | immunogenicity of COVID-19 vaccination |
| NCT04884685 | 07/05/2021 | Not recruiting | China | 3-17 | 500 | Interventional  Phase 2 | SARS-CoV-2 Inactivated Vaccine; | Safety index-incidence of ARs |
| NCT04869592 | 24/04/2021 | Recruiting | China | ≥3 | 3580 | Interventional  Phase 1/2 | Recombinant SARS-CoV-2 Vaccine (CHO cell) | the incidence and severity of any ARs/AEs within 30 minutes after each dose of vaccination; the incidence of abnormal blood biochemistry, blood routine, blood coagulation function and urine routine on the 4th day after each dose of vaccination; the incidence and severity of ARs/AEs within 0-7 days after each dose of vaccination; the incidence and severity of non-collective ARs/AEs within 30 days after each dose of vaccination; the incidence and severity of AEs leading to withdrawal within 30 days after each dose of vaccination; the incidence of SAE from the first dose of vaccination to 12 months after the full course of vaccination; the incidence of AESI from the first dose of vaccination to 12 months after the full course of vaccination; GMT of SARS-CoV-2 specific neutralizing antibody (wild Strains) on the 15th day after the full course of vaccination; the incidence and severity of any ARs/AEs within 30 minutes after each dose of vaccination; the incidence and severity of ARs/AEs within 0-7 days after each dose of vaccination; the incidence and severity of non-collective ARs/AEs within 30 days after each dose of vaccination; the incidence and severity of non-collective ARs/AEs within 40 days after each dose of vaccination; the incidence and severity of non-collective ARs/AEs within 60 days after each dose of vaccination; the incidence and severity of AEs leading to withdrawal within 30 days after each dose of vaccination; the incidence and severity of AEs leading to withdrawal within 40 days after each dose of vaccination; the incidence and severity of AEs leading to withdrawal within 60 days after each dose of vaccination; the incidence of SAE from the first dose of vaccination to 12 months after the full course of vaccination; the incidence of AESI from the the first dose of vaccination to 12 months after the full course of vaccination; GMT of SARS-CoV-2 specific neutralizing antibody (wild Strains) on the 15th day after the full course of vaccination |
| NCT04865900 | 28/04/2021 | Recruiting | Israel | 16-60 | 500 | Observational | Diagnostic Test: Laboratory Blood Tests; Diagnostic Test: ECG; Diagnostic Test: PCR; Diagnostic Test: Antibody test; Diagnostic Test: Echocardiography; Diagnostic Test: MRI | Elevated Troponin |
| NCT04863638 | 27/04/2021 | Recruiting | China | ≥3 | 4400 | Interventional  Phase 4 | COVID-19 Vaccine (Vero Cell), Inactivated; | GMT of anti-SARS-CoV-2 neutralizing antibody; The four-fold increase rate of anti-SARS-CoV-2 neutralizing antibody |
| NCT04848584 | 30/03/2021 | Not recruiting | NR | ≥16 | 999 | Observational | Pfizer-BioNTech COVID-19 Vaccine | The effectiveness of 2 doses of BNT162b2 (i.e., fully vaccinated) against hospitalization for ARI due to SARS-CoV-2 infection |
| NCT04832932 | 02/04/2021 | Recruiting | United States; Kenya; United Kingdom; | ≥16 | 1000 | Observational | COVID-19 vaccine | ARs/AEs |
| NCT04816643 | 19/03/2021 | Recruiting | United States; Finland; Poland; Spain | 6m-11y | 4644 | Interventional  Phase 1/2 | BNT162b2 | Percentage of participants in Phase 1 reporting local reaction in each dose level in each age group; Percentage of participants in Phase 1 reporting systemic events in each dose level in each age group; Percentage of participants in Phase 1 reporting AEs in each dose level in each age group; Percentage of participants in Phase 1 reporting SAEs in each dose level in each age group; Percentage of participants in Phase 2/3 reporting local reaction in each dose level in each age group; Percentage of participants in Phase 2/3 reporting systemic events in each dose level in each age group; Percentage of participants in Phase 2/3 reporting AEs in each dose level in each age group; Percentage of participants in Phase 2/3 reporting SAEs in each dose level in each age group; In Phase 2/3 participants, GMR of SARS-CoV-2 neutralizing titers in participants in each age group at selected dose level to those 16 to 25 years of age in study C4591001 |
| NCT04811391 | 19/03/2021 | Recruiting | Albania | ≥16 | 1500 | Observational | COVID-19 vaccine | COVID-19 vaccine effectiveness; COVID-19 PCR confirmation |
| NCT04800133 | 01/03/2021 | Recruiting | China | 11-100 | 900 | Interventional  Phase 2 | Tozinameran; Oxford-AstraZeneca COVID-19 vaccine; CoronaVac | ARs/AEs; Binding antibody response on ELISA; Neutralizing antibody response on PRNA; T cell response on flow cytometry |
| NCT04796896 | 11/03/2021 | Recruiting | United States | 6m-11y | 6750 | Interventional  Phase 2/3 | mRNA-1273 | Number of Participants with Solicited Local and Systemic ARs; Number of Participants with Unsolicited AEs; Number of Participants with MAAEs; Number of Participants with SAEs; Number of Participants with AEs of Special Interest, including Multisystem Inflammatory Syndrome in Children (MIS-C);Number of Participants with Serum Antibody (Ab) Levels that Meet or Exceed the Threshold of Protection from COVID-19; GM of SARS-CoV-2 Specific Neutralizing Antibody (nAb); Seroresponse Rate of Vaccine Recipients |
| NCT04773067 | 23/02/2021 | Recruiting | China (Taiwan) | 12-85 | 3850 | Interventional  Phase 2 | UB-612 | GMT of SARS-CoV-2 neutralizing antibody; Seroconversion rate (SCR) of SARS-CoV-2 neutralizing antibody; Safety evaluation |
| NCT04761822 | 17/02/2021 | Recruiting | United States | 12-69 | 3400 | Interventional  Phase 2 | Moderna COVID-19 Vaccine; Pfizer-BioNTech COVID-19 Vaccine | Proportion of participants who experience a systemic allergic reaction to either dose of the Pfizer-BioNTech COVID-19 Vaccine; Proportion of participants who experience a systemic allergic reaction to either dose of the Moderna COVID-19 Vaccine |
| NCT04713553 | 15/01/2021 | Recruiting | United States | 12-50 | 1530 | Interventional  Phase 3 | BNT162b2 | GMR of SARS-CoV-2 full-length S-binding antibody levels between US lots (Arms 1, 2 and 3) in participants without evidence of infection during the study; GMR of SARS-CoV-2 full-length S-binding antibody levels between the EU lot (Arm 4) and pooled US lots (Arms 1, 2, and 3) in participants without evidence of infection during the study; GMR of SARS-CoV-2 neutralizing antibody levels between the 20-microgram dose group (Arm 5) and the corresponding 30-microgram dose group (Arm 1, 2, or 3) in participants without evidence of SARS-C0V-2 infection during the study.; Percentage of participants reporting local reactions; Percentage of participants reporting systemic events; Percentage of participants reporting AEs; Percentage of participants reporting SAEs; GMT of SARS-CoV-2 reference strain and B.1.351 strain neutralizing antibody levels for Booster Arm 1 (BNT162b2) and Booster Arm 2 (BNT162b2.B.1.351).;Geometric Mean IgG Concentrations (GMC) of SARS-CoV-2 full-length S-binding antibody levels for Booster Arm 1 (BNT162b2) and Booster Arm 2 (BNT162b2.B.1.351).; GMFR of SARS-CoV-2 reference strain and B.1.351 strain neutralizing antibody levels for Booster Arm 1 (BNT162b2) and Booster Arm 2 (BNT162b2.B.1.351).; GMFR of SARS-CoV-2 full-length S-binding antibody levels for Booster Arm 1 (BNT162b2) and Booster Arm 2 (BNT162b2.B.1.351).;Percentages of participants with seroresponse (based on neutralizing titers) to the reference strain.; Percentages of participants with seroresponse (based on neutralizing titers) to the B.1.351 strain. |
| NCT04649151 | 30/11/2020 | Not recruiting | United States | 12-17 | 3732 | Interventional  Phase 2/3 | mRNA-1273 | Number of Participants with Solicited Local and Systemic ARs; Number of Participants with Unsolicited AEs; Number of Participants with SAEs, MAAEs, or AEs of Special Interest of Multisystem Inflammatory Syndrome in Children (MIS-C); Number of Participants With Serum Antibody (Ab) Levels that Meet or Exceed the Threshold of Protection From COVID-19; GM of the Serum Ab Level; Seroresponse Rate of Vaccine Recipients |
| NCT04611802 | 30/10/2020 | Recruiting | United States; Mexico; Puerto Rico; | ≥12 | 33000 | Interventional  Phase 3 | SARS-CoV-2 rS/Matrix-M1 Adjuvant | Pediatric Expansion: Incidence and Severity of SAEs from Month 12 to Month 24;Pediatric Expansion: Incidence and Severity of MAAEs Attributed to Study Vaccine from Month 12 to Month 24;Pediatric Expansion: Incidence and Severity of AESIs from Month 12 to Month 24;Pediatric Expansion: Antibodies to SARS-CoV-2 Nucleoprotein (NP) at Specific Time Points; Pediatric Expansion: Deaths Due to Any Cause; Main Study and Pediatric Expansion: Participants with Symptomatic Mild, Moderate, or Severe Coronavirus Disease 2019 (COVID-19);Pediatric Expansion: Reactogenicity Incidence and Severity; Pediatric Expansion: Incidence and Severity of MAAEs Through Day 49;Pediatric Expansion: Incidence and Severity of Unsolicited AEs Through Day 49;Pediatric Expansion: Incidence and Severity of MAAEs Attributed to Study Vaccine Through Month 12;Pediatric Expansion: Incidence and Severity of SAEs Through Month 12;Pediatric Expansion: Incidence and Severity of AEs of Special Interest Through Month 12 |
| NCT04566770 | 22/09/2020 | Recruiting | China | ≥6 | 481 | Interventional  Phase 2 | Recombinant Novel Coronavirus Vaccine (Adenovirus Type 5 Vector); | Safety indexes of ARs; Immunogenicity indexes of GMT; Immunogenicity indexes of neutralizing antibody |
| NCT04551547 | 15/09/2020 | Recruiting | China | 3-17 | 552 | Interventional  Phase 1/2 | inactivated SARS-CoV-2 vaccine a | Safety index-incidence of ARs; Immunogenicity index-seroconversion rates of neutralizing antibody |
| NCT04528719 | 20/08/2020 | Recruiting | United States | 12m-79y | 620 | Interventional  Phase 1 | mRNA-1345 | Number of Participants with Solicited Local and Systemic ARs; Number of Participants with Unsolicited AEs; Number of Participants with Serious AEs or MAAEs |
| NCT04471519 | 11/07/2020 | Not recruiting | India | 12-65 | 755 | Interventional  Phase 1/2 | BBV152 | Frequency of Grade 1 or higher solicited AEs; Frequency of Grade 2 or higher lower respiratory infections (LRI); Frequency of SAEs; Percentage of vaccinees with a =4-fold rise in serum RSV-neutralizing antibody titer; Peak titer of vaccine virus shed; Proportion of vaccinees infected with vaccine virus in Group 1 |
| NCT04368728 | 27/04/2020 | Recruiting | United States; Argentina; Brazil; Germany; South Africa; Turkey | ≥12 | 43998 | Interventional  Phase 2/3 | BNT162b1; BNT162b2; BNT162b2SA | Phase 1: Occurrence of AEs and SAEs  Phase 2: Evaluation of Neutralizing Antibody Titers |
| NCT04299724 | 05/03/2020 | Recruiting | China | 6m-80y | 100 | Interventional  Phase 1 | Pathogen-specific aAPC | Percentage of participants in Phase 1 reporting local reactions; Percentage of participants in Phase 1 reporting systemic events; Percentage of participants in Phase 1 reporting AEs; Percentage of participants in Phase 1 reporting SAEs; Percentage of Phase 1 participants with abnormal hematology and chemistry laboratory values; Percentage of Phase 1 participants with abnormal hematology and chemistry laboratory values; Percentage of Phase 1 participants with abnormal hematology and chemistry laboratory values; Percentage of Phase 1 participants with grading shifts in hematology and chemistry laboratory assessments; Percentage of Phase 1 participants with grading shifts in hematology and chemistry laboratory assessments; Percentage of Phase 1 participants with grading shifts in hematology and chemistry laboratory assessments; In the first 360 participants randomized into Phase 2/3, percentage of participants reporting local reactions; In the first 360 participants randomized into Phase 2/3, percentage of participants reporting systemic events; In the first 360 participants randomized into Phase 2/3, percentage of participants reporting AEs; In the first 360 participants randomized into Phase 2/3, percentage of participants reporting SAEs; In a subset of at least 6000 participants randomized in Phase 2/3, percentage of participants reporting local reactions; In a subset of at least 6000 participants randomized in Phase 2/3, percentage of participants reporting systemic events; Percentage of participants in Phase 2/3 reporting AEs; Percentage of participants in Phase 2/3 reporting SAEs; Confirmed COVID-19 in Phase 2/3 participants without evidence of infection before vaccination; Confirmed COVID-19 in Phase 2/3 participants with and without evidence of infection before vaccination; Percentage of participants 12-15 years of age in Phase 3 reporting AEs; Percentage of participants 12-15 years of age in Phase 3 reporting AEs; In participants 12-15 years of age randomized in Phase 3, percentage of participants reporting local reactions; In participants 12-15 years of age randomized in Phase 3, percentage of participants reporting systemic events; In participants who receive BNT162b2SA given as 1 or 2 doses, percentage of participants reporting AEs; In participants who receive BNT162b2SA given as 1 or 2 doses, percentage of participants reporting SAEs; In participants, who receive BNT162b2SA given as 1 or 2 doses, percentage of participants reporting local reactions; In participants who receive BNT162b2SA given as 1 or 2 doses, percentage of participants reporting systemic events; In participants who receive a third dose of BNT162b2, percentage of participants reporting AEs; In participants who receive a third dose of BNT162b2, percentage of participants reporting SAEs; In participants who receive a third dose of BNT162b2, percentage of participants reporting local reactions; In participants who receive a third dose of BNT162b2, percentage of participants reporting systemic events; Noninferiority of the SARS-CoV-2 reference strain neutralizing titers after a third dose of BNT162b2 at 30 g compared to after 2 doses of BNT162b2, in the same individuals; Noninferiority of the SARS-CoV-2 SA strain neutralizing titers after one dose of BNT162b2SA compared to the SARS-CoV-2 reference strain neutralizing titers after 2 doses of BNT162b2, in the same individuals; Noninferiority of the SARS-CoV-2 SA strain neutralizing titers after 2 doses of BNT162b2SA compared to the SARS-CoV-2 reference strain neutralizing titers after 2 doses of BNT162b2 |
| NCT04276896 | 17/02/2020 | Recruiting | China | 6m-80y | 100 | Interventional  Phase 1/2 | LV-SMENP-DC vaccine | Frequency of vaccine events; Frequency of serious vaccine events; Proportion of subjects with positive T cell response |

GMT: The Geometric Mean Titer; AEs: Adverse Events; SAEs: Serious Adverse Events; MAAEs: Medically Attended Adverse Events; Ars: adverse reactions; GMFR: Geometric Mean Fold Rise; GM: Geometric Mean; GMR: Geometric Mean Ratio; N/A: Not Applicable
